# Amygdala-related electrical fingerprint is modulated with neurofeedback training and correlates with deep-brain activation: Proof-of-concept in borderline personality disorder

**DOI:** 10.1101/2023.03.28.23287782

**Authors:** Malte Zopfs, Miroslava Jindrová, Guy Gurevitch, Jackob N. Keynan, Talma Hendler, Sarah Baumeister, Pascal-M. Aggensteiner, Sven Cornelisse, Daniel Brandeis, Christian Schmahl, Christian Paret

## Abstract

**Background:** The modulation of brain circuits of emotion is a promising pathway to treat Borderline Personality Disorder (BPD). Precise and scalable approaches have yet to be established. Two studies investigating the Amygdala-related Electrical Fingerprint (Amyg-EFP) in BPD are presented: One study addressing the deep-brain correlates of Amyg-EFP, and a second study investigating neurofeedback (NF) as a means to improve brain self-regulation.

**Methods:** Study 1 combined EEG and simultaneous fMRI to investigate the replicability of Amyg-EFP-related brain activation found in the reference dataset (N=24 healthy subjects, 8 female; re-analysis of published data) in the replication dataset (N=16 female individuals with BPD). In the replication dataset, we additionally explored how the Amyg-EFP would map to neural circuits defined by the Research Domain Criteria. Study 2 investigated a 10-session Amyg-EFP NF training in parallel to a 12-weeks residential Dialectical Behavior Therapy (DBT) program. N=15 patients with BPD completed the training, N=15 matched patients served as DBT-only controls.

**Results:** Study 1 replicated previous findings and showed significant amygdala BOLD-activation in a whole-brain regression analysis with the Amyg-EFP. Neurocircuitry activation (negative affect, salience, and cognitive control) was correlated with the Amyg-EFP signal. Study 2 showed significant learning of Amyg-EFP modulation with NF training. No clinical benefits of NF beyond DBT-only were observed.

**Conclusions:** Recorded via scalp EEG, the Amyg-EFP picks up brain activation of high relevance for emotion. Administering Amyg-EFP NF in addition to standardized BPD treatment was shown to be feasible. Clinical utility remains to be investigated.

## 1 Introduction

The modulation of deep brain regions is a promising treatment option for various mental disorders. One such disorder is Borderline Personality Disorder (BPD), which is characterized by pervasive emotion dysregulation related to aberrations in the amygdala and in prefrontal-limbic networks (Schulze et al., 2016; Sicorello & Schmahl, 2021). To date, little evidence exists whether those with BPD can benefit from neurofeedback (NF) (Howard et al., 2013; Paret et al., 2016; Zaehringer et al., 2019). In principle, NF allows individuals to self-modulate their brain activation: via a brain-computer interface, patients observe and control their brain activation in real-time (Paret & Hendler, 2020). The amygdala lends itself as a target for NF in patients with BPD. However, generating precise feedback from deep-brain regions, such as the amygdala, requires cost-intensive brain scanning with functional Magnetic Resonance Imaging (fMRI) (Paret et al., 2019). The limited availability of MR machines, the cost of using them and the aversion of many patients against lengthy MRI measurements represent considerable barriers for amygdala-NF studies. To overcome these disadvantages of fMRI, an electrocortical surrogate of deep-brain activation was developed: The amygdala-related Electrical Fingerprint (Amyg-EFP) (Meir-Hasson et al., 2016). The Amyg-EFP can be used as a proxy for deep brain activation, thereby allowing for amygdala-NF training outside of an MR machine. Previously, Amyg-EFP NF was found to be effective in ameliorating symptoms of post-traumatic stress (Fruchtman-Steinbok et al., 2021; Keynan et al., 2019). The present work had two main goals: (1) To assess whether the Amyg-EFP can be used to probe deep-brain activation in BPD, and (2) to investigate the feasibility of Amyg-EFP NF training with patients who are undergoing a residential Dialectical Behavior Therapy (DBT) program, a standard treatment of BPD (Bohus et al., 2021).

Measuring subcortical brain activity via electroencephalography (EEG) is an intricate problem, in particular when it comes to deep-brain structures. To overcome the limited anatomical specificity of EEG, the Amyg-EFP has been developed based on simultaneously acquired fMRI and EEG. The algorithm underlying the computation of the Amyg-EFP uses time and frequency information from band-widths recorded with three scalp EEG-electrodes: ground, reference and one more electrode (Meir-Hasson et al., 2016). The resulting signal is an EEG surrogate of Blood Oxygenation Level Dependent (BOLD) activation, optimized for the amygdala. Feedback based on this signal is supposed to facilitate NF training to regulate amygdala activation.

In order to transfer this technology to different labs and clinical centers, and to introduce Amyg-EFP to the treatment of a different clinical population, we aimed to investigate whether previous results were replicable and whether they would generalize to BPD. We performed two studies: For study 1, we recorded simultaneous EEG-fMRI data from patients with BPD to test whether (A) the Amyg-EFP would predict amygdala activation, and (B) whether the fMRI-BOLD pattern prediction would be consistent with benchmark findings from Keynan et al. (2016), i.e., whether the results would replicate and generalize to BPD. Study 2 assessed the feasibility of Amyg-EFP NF training within the context of residential DBT and compared outcomes to a DBT-only control group not receiving NF.

We hypothesized, first, that the Amyg-EFP signal would predict amygdala-BOLD activation, and second, that effects from previous research could be replicated with a new dataset. That is, we assumed that the whole-brain pattern of effect sizes from a ‘reference dataset’ would not be different from the pattern observed in the BPD dataset. Additionally, we explored whether patients improve regulation of the Amyg-EFP with NF training. Lastly, we assessed the feasibility of NF in the context of residential DBT.

## 2 Methods and Materials

### 2.1 Replication of Amyg-EFP-related brain pattern in Borderline Personality Disorder

We analyzed a new dataset recorded in an adolescent BPD-sample (the ‘replication dataset’) and compared it to a ‘reference dataset’ (Keynan et al., 2016) to assess two facets of replicability: 1) Replication of significant amygdala activation, and 2) replication of effect sizes (ES).

#### Description of the reference dataset

N=24 healthy participants (age (mean/SD): 26.75/3.81 years, 8 females) underwent a simultaneous EEG-fMRI measurement, which was the last session of a multi-session experiment. The study also included 4 EEG-only NF sessions and a pre-fMRI assessment, which were not analyzed for this paper. Participants completed five functional runs (baseline, NF run 1-4) and one anatomical brain scan. The experimental group (N=17) received feedback from the Amyg-EFP and the control group (N=7) from the alpha-theta ratio. More details can be found in the original publication (Keynan et al., 2016). During NF, participants saw an animation of a skateboarder, who gained in speed when Amyg-EFP activation increased. In each of the 4 NF runs, they had to decrease the speed of the skateboarder by modulating their brain activity. NF blocks (60 s) alternated with Rest (60 s) and finger-tapping blocks (30 s).

##### EEG data acquisition

EEG was acquired with an MR-compatible BrainAmp-MR amplifier (BrainProducts, Munich, Germany) and BrainCap electrode cap with sintered Ag/AgCl ring electrodes (30 channels, 1 ECG channel, 1 EOG channel; Falk Minow Services, Herrsching-Breitbrunn, Germany).

Electrodes were positioned to 10/20 system with the reference electrode between FCz and Cz. The raw EEG was sampled at 250 Hz and recorded using Brain Vision Recorder software (Brain Products).

##### Online Calculation of Amyg-EFP amplitude

The Amy-EFP signal was calculated online from raw EEG data using a built-in automated average artifact subtraction method implemented in BrainVision RecView (BrainProducts). RecView was custom modified to enable export of the corrected EEG data in real time through a TCP/IP socket. Preprocessing algorithm and EFP calculation models were compiled from MATLAB R2009b to Microsoft.NET in order to execute it within the BrainVision RecView EEG Recorder system. Data was then marshaled to a MATLAB.NET compiled dll that calculated the value of the EFP amplitude every 3 seconds. The online generated EFP data were used for analyses.

##### fMRI data acquisition

Structural and functional scans were performed using a GE 3T Signa Excite echo speed scanner with an 8-channel head coil, and a resonant gradient echoplanar imaging system. The scanner was located at the Wohl Institute for Advanced Imaging at the Tel-Aviv Sourasky Medical Center. A T1-weighted 3D axial spoiled gradient echo pulse sequence (TR/TE = 7.92/2.98 ms, flip angle = 15º, pixel size = 1 mm, FOV = 256 × 256 mm, slice thickness = 1 mm) was applied to provide high-resolution structural images. Functional whole-brain scans were performed in an interleaved top-to-bottom order, using a T2*-weighted gradient echo planar imaging pulse sequence (TR/TE = 3000/35 ms, flip angle = 90º, pixel size = 1.56 mm, FOV = 200 × 200 mm, slice thickness = 3 mm, 39 slices per volume).

##### fMRI preprocessing and analysis

fMRI data was imported to the Brain Imaging Data Structure (BIDS) (Gorgolewski et al., 2016), using adapted code-scripts based on the Rapid, automated BIDS conversion (RaBIDS) pipeline (Paret, 2023b), and preprocessed with fMRIPrep v20.0.6 ((Esteban et al., 2019), see Supplement). SPM12 v7771 (The Wellcome Centre for Human Neuroimaging, London UK) was used for first-level analysis. The initial 4 volumes were discarded to allow longitudinal magnetization to reach equilibrium. The General Linear Model (GLM) for the first-level analysis contained 7 orthogonalized predictors: The Amyg-EFP time-course, i.e., the effect of interest, and the 6 realignment regressors for nuisance regression. The Amyg-EFP timecourse was not convolved with the Haemodynamic Response Function (HRF). No high-pass filter was applied to the data. SPM’s autoregression AR(1) model was applied. Data quality was assessed based on fMRIPrep’s html-output files. To exclude spurious effects, data coinciding with large movements (framewise displacement (FD)>4 mm) was excluded from the analysis (either full runs, or initial/final volumes if movements happened in the beginning/end of the scan). This affected three runs in total.

#### Replication in sample with Borderline Personality Disorder

Patients were eligible to participate in this study during the first half of the residential DBT-program (i.e., 6 weeks) if they were female, fulfilled 4 or more DSM-IV BPD criteria as determined by a trained clinician, and were aged 18-25 years. They were excluded in case of pharmacotherapy with benzodiazepines, pregnancy, epilepsy, traumatic brain injury, brain tumor or otherwise severe neurological or medical history, BMI > 16.5, and if they fulfilled the usual MRI exclusion criteria. Participants had to be abstinent from illicit drugs and alcohol. The resulting replication sample consisted of N=16 participants (21.3/2.19 years, see Supplementary Table S1) The simultaneous EEG-MR-scan was composed of 3 runs: a resting-state scan (6 min), a short NF run (6 min) and a long NF run (22 min), during which individuals received fMRI-NF. Participants were instructed to downregulate a visual analogue scale illustrating brain activity (short NF run) or to increase and decrease brain activity in alternating blocks (long NF run, Supplement Figure S1).

##### EEG data acquisition

The EEG was recorded during image acquisition inside the scanner using an MRI-compatible EEG system with a 5 kHz sampling rate, 32 mV input range and 0.1–250 Hz band-pass filters. The signal was recorded by equidistantly spaced sintered silver/silverchloride (Ag/AgCl) scalp electrodes using EEG caps with twisted and fixed electrode cables (64Ch BrainCap-MR with Multitrodes; Easycap, Munich, Germany). The 64-channel EEG montage included most 10–10 system positions. Fz served as recording reference, and AFz as the ground electrode. Four additional electrodes were placed to record the electro-oculogram (EOG) and the electrocardiogram (ECG). The signal was transmitted from two MRI-compatible amplifiers (BrainAmp MR, BrainProducts, Gilching, Germany) outside the scanner via optic fibers. Electrode impedances were kept below 20 kΩ, except for ECG and EOG electrodes (<30 kΩ) as well as reference and ground (<10 kV). The quality of the EEG was assessed during the MR-scan, using online correction software (RecView BrainProducts, Gilching, Germany).

##### fMRI data acquisition

Structural and functional scans were performed using a 3 Tesla MRI Scanner (Trio, Siemens Medical Solutions, Erlangen, Germany) with a 20-channel head coil. After the first 8 study subjects the MR-scannerreceived an upgrade (PRISMAfit, Siemens Medical Solutions, Erlangen, Germany) with which all remaining subjects of the study were scanned. Functional images of the BOLD contrast were acquired with a gradient echo T2*-weighted echo-planar-imaging sequence (TE= 30ms, TR= 2s, FOV: 192×192mm, flip angle= 80°, inplane resolution= 3×3mm). One volume comprised 36 slices tilted −20° from AC-PC orientation with a thickness of 3mm and a slice gap of 1 mm. Participants had their heads lightly restrained in the coil using soft pads. The resting-state scan comprised 180 volumes each, while the experimental runs for the 6min down-regulation NF were 186 volumes each and the 22min up-down-regulation NF 666 volumes each. T1-weighted anatomical images were acquired with a Magnetization Prepared Rapid Acquisition Gradient Echo sequence (TE= 3.03ms, TR= 2.3 s, 192 slices and FOV= 256×256mm).

##### fMRI preprocessing and analysis

Preprocessing and analysis steps were identical with the analysis of the reference dataset. Heavily movement-affected volumes were repaired, using the ArtRepair toolbox (https://cibsr.stanford.edu/tools/human-brain-project/artrepair-software.html). After re-estimating the SPM model using the repaired volumes, the improvement between the repaired and the original model was assessed based on global quality estimates (whole-brain contrast-to-noise ratio). The re-estimated SPM model from 4 subjects was used for further analysis, as quality improved >5% relative to the original SPM model.

##### Replication of effect sizes

Following Gerchen et al. (2021), we tested whether the effect sizes found in Keynan and coworkers’ (2016) ‘reference sample’ would fall into the 90% Confidence Interval (CI) of our ‘replication sample’. We assessed only positive effects (i.e., Hedge’s g>0), resulting in a one-sided significance threshold for replication of p<0.05. The statistical analysis was preregistered before results were known (https://doi.org/10.17605/OSF.IO/KYCR6).

#### Mapping Amyg-EFP signal to neurocircuitries and nuisance signal

We analyzed correlations of the Amyg-EFP signal with BOLD-signal time courses from 5 neurocircuitries, which were defined according to masks provided by Goldstein-Piekarski et al. (2022): Negative affect neurocircuitry, salience neurocircuitry, default-mode network (DMN), cognitive control neurocircuitry, and positive affect neurocircuitry. Additionally, we analyzed correlations with the BOLD signal time course from sensory-motor domains. Visual cortex was defined as area hOc1, auditory cortex as area TE 1, and motor cortex as areas 4a and 4p (Eickhoff et al., 2005). We used SPM’s VOI tool to extract the eigenvariate from each region, which was then adjusted for the effect of interest, i.e., the F-contrast received from the Amyg-EFP predictor.

### 2.2 Feasibility of neurofeedback training

The study design and results are reported according to best-practice guidelines and provide the CRED-nf checklist (Ros et al., 2020) in the Online Supplement [*to reviewers/editor*: *will be provided before publication in peer-reviewed journal*].

#### Sample

N=29 female patients diagnosed with BPD were allocated to the NF group. N=15 of them received the full dose of ten Amyg-EFP NF sessions over five weeks in addition to their residential DBT treatment. N=22 female patients diagnosed with BPD were assigned to a control group, receiving no NF training in addition to DBT treatment (no-NF group). N=15 of them completed the study. Eligibility criteria were the same as reported above. The NF group (N=15, age: 21.4/1.84 years) did not differ in age from the no-NF group (N=15, 20.7/1.98; T(27.86)=-1.046, p=0.305). The two groups did not differ in clinical characteristics such as psychopathology (BSL-23, (Wolf et al., 2009)), comorbidities or psychotropic medication (Table 1). No differences were observed between the groups in baseline depression (BDI-II, (Steer et al., 1999)), anxiety (STAI, (Laux et al., 1981)), affective lability (ALS, (Harvey et al., 1989)) and alexithymia scores (TAS-26, (Taylor et al., 1985), Tables 1, 2). Supplemantary Figure S2 presents a comprehensive patient flow chart.

**Table 1.**
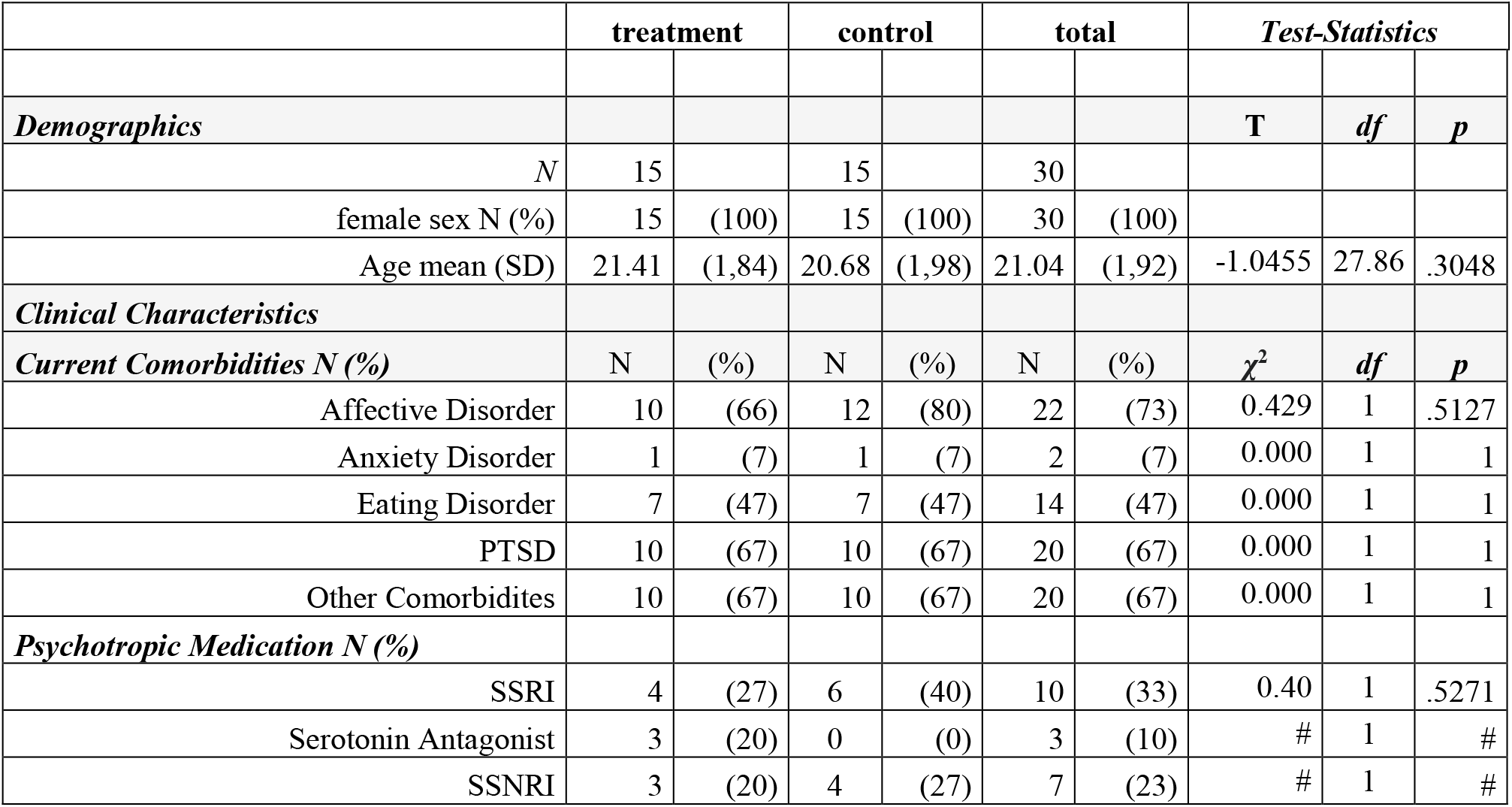
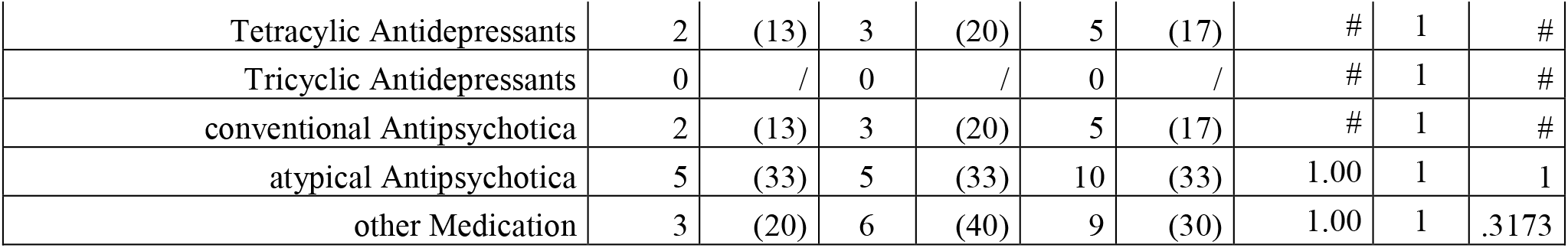
Study 2 sample characteristics: demographics and psychiatric characteristics.

**Table 2.** Study 2 sample characteristics: clinical psychological characteristics

| *Self-report measures* | **treatment** |  | **control** |  | **total** |  | *Test-Statistics* |  |  |
| --- | --- | --- | --- | --- | --- | --- | --- | --- | --- |
| *N* | 15 |  | 14 |  | 29 |  |  |  |  |
| *Beck Depression Inventory (BDI)* | Mean | SD | Mean | SD | Mean | SD | **T** | **df** | **p** |
| Total | 38.80 | 8.51 | 38.64 | 13.15 | 38.72 | 10.79 | -0.038 | 27 | .970 |
| *State Trait Anxiety Inventory (STAI)* |  |  |  |  |  |  |  |  |  |
| Total | 65.67 | 6.41 | 63.43 | 8.60 | 64.59 | 7.50 | -0.798 | 27 | .432 |
| *Affect Liability Scale (ALS)* |  |  |  |  |  |  |  |  |  |
| Total | 97.27 | 24.76 | 99.79 | 31.60 | 98.48 | 27.78 | 0.240 | 27 | .812 |
| Depression | 21.53 | 4.52 | 23.07 | 5.73 | 22.28 | 5.11 | 0.806 | 27 | .427 |
| Hypomania | 17.33 | 7.88 | 18.14 | 9.22 | 17.72 | 8.41 | 0.255 | 27 | .801 |
| Biphasic shifts | 15.87 | 5.59 | 16.00 | 6.30 | 15.93 | 5.84 | 0.060 | 27 | .952 |
| Anxiety | 12.47 | 3.27 | 13.21 | 4.58 | 12.83 | 3.90 | 0.509 | 27 | .615 |
| Anger | 10.73 | 5.57 | 9.93 | 6.83 | 10.34 | 6.11 | 0.349 | 27 | .730 |
| Anxiety Depression | 19.33 | 3.37 | 19.43 | 5.23 | 19.38 | 4.29 | 0.059 | 27 | .954 |
| *Toronto Alexithymia Scale (TAS-26)* |  |  |  |  |  |  |  |  |  |
| Total | 58.20 | 7.84 | 62.64 | 7.19 | 60.34 | 7.73 | 1.587 | 27 | .124 |
| Identification of one's feelings | 24.47 | 5.73 | 26.71 | 4.20 | 25.55 | 5.09 | 1.198 | 27 | .241 |
| Difficulty Describing Feelings | 20.20 | 3.61 | 19.86 | 3.11 | 20.03 | 3.32 | -0.273 | 27 | .787 |
| External thinking | 13.53 | 3.62 | 16.07 | 4.39 | 14.76 | 4.15 | 1.702 | 27 | .100 |
| *N* | 15 |  | 15 |  | 30 |  |  |  |  |
| **Borderline Symptom List (BSL)** | 2.28 | 0.85 | 2.51 | 0.73 | 2.39 | 0.80 | 0.759 | 27 | .454 |
| during the week of study inclusion |  |  |  |  |  |  |  |  |  |

*Note: SD, standard deviation; SSRI, Selective serotonin reuptake inhibitor; SNRI, Serotonin–norepinephrine reuptake inhibitor; #, frequencies too small for test; †p<.10; \*p<.05; \*\*p<.01,*

#### General procedure

Participants for this study were recruited in the inpatient units of the Department of Psychosomatic Medicine and Psychotherapy, Central Institute of Mental Health, between May 2018 and February 2021. We approached patients during their first 3 weeks of the 12-weeks DBT program. The control arm was recruited after completion of the NF group.

Questionnaires and a MR scan were completed at the beginning of the study (pre-measurement), followed by 10 EEG-NF-trainings over the course of 5 weeks for the NF group. After 5 weeks, the questionnaires and MR scan were completed again by both groups (post-measurement).

The authors assert that all procedures contributing to this work comply with the ethical standards of the relevant national and institutional committees on human experimentation and with the Helsinki Declaration of 1975, as revised in 2008. The experiments were conducted at the CIMH in Mannheim, Germany. All participants provided informed written consent before participation and received no reimbursement for participation. Two subjects who were recruited for the simultaneous EEG-fMRI scan only, and did not take part in any other part of the study, received 50€.

#### EEG acquisition for neurofeedback training

EEG was recorded with 3 electrodes: the ground (AFz), reference (FCz) and active electrode (Pz) were mounted according to the 10-10 system using a standardized cap (Easycap, Herrsching, Germany). The EEG-signal was recorded with BrainAmp ExG-amplifier (BrainProducts, Gilching, Germany) with a sampling rate of 250Hz and the following filters: Low-Cutoff= 3Hz, High-Cutoff= 70Hz and no Notch filter. Electrode impedances were kept below 5kΩ. The recording software was BrainVision Recorder (BrainProducts, Gilching, Germany).

#### Feedback protocol

Participants were sitting with eyes open in a relaxed position in front of a black computer screen. A piano melody of 3 seconds was repeatedly played to participants (Kinreich et al., 2014). Participants were instructed to downregulate the volume. The study was designed to assess the effects of Amyg-EFP downregulation and was preregistered accordingly^1^. Due to a programming error that was revealed after the completion of data acquisition, the audio-volume was inversely coupled with Amyg-EFP activation. That is, the piano volume decreased when patients upregulated Amyg-EFP.

Each of the 10 training sessions lasted 20 minutes and consisted of 5 cycles with a duration of 4 min each. Every cycle was composed of a baseline block of 1 minute followed by a feedback block of 3 minutes. The audio volume was adjusted to the measured Amyg-EFP signal in feedback blocks and was fixed at 70% of the maximum volume in baseline blocks. Participants reached the minimum/maximum volume when the Amyg-EFP signal was < −2 SD/>2SD from the preceding baseline mean (baseline values of the initial 6s were dropped).

#### Clinical self-report outcomes

Self-report and training success data were analyzed using R software version 4.2.2. Clinical questionnaires were assessed 2-7 days before the first measurement to ensure matching of groups at baseline (‘pre’). Clinical outcomes (BDI, ALS, TAS-26) were assessed again 2-7 days after the last NF session (‘post’), or in case of the no-NF group, 5 weeks after the pre-measurement. Extreme values were defined as values x < Q (quartile) 1 - 3*IQR (interquartile range) or x > Q3 + 3*IQR according to the rstatix package (Kassambara, 2023a) and were excluded from analysis. The afex package was used to analyze mixed ANOVAs (Singmann et al., 2023).

#### Neurofeedback training success

Training success was quantified as the personal effect size (PES) (Paret et al., 2019). PES measures the change of the Amyg-EFP signal from a NF block (3 min; 60 samples) relative to the preceding baseline block (1 min; 20 samples) divided by the pooled standard deviation.

PES values of each block were averaged and analyzed with multilevel regression analysis using the lme4 package (Bates et al., 2015). The model reflected the nested data structure of blocks within sessions and sessions within participants, and included a Subject random effect. To analyze the linear effect of Session (i.e., assuming an incremental learning effect across training), Session was included as a random effect. The random effect for the “Subject x Session” interaction was included. The fixed effect “Session” was assessed for significance. Session was centered on the first run (i.e., *x centered* = *x* − 1). Data was assessed for heteroscedasticity via visual inspection of quantile-quantile plots.

#### Correlation of self-report with neurofeedback success

The final 2 sessions and the initial 2 sessions were averaged and the difference was calculated. Correlations were calculated with the change in clinical measures (i.e., post minus pre). The R package ggpubr was used to assess explained variance R and significance p of pearson correlation (Kassambara, 2023b).

### 2.3 Availability of materials

Self-report data, individual fMRI and EEG data is available on reasonable request and can be shared in line with applicable data protection regulations. Analysis code of self-report and NF training data is openly available (Paret, 2023a). The T-map from the second-level fMRI analysis (i.e., aggregated data across subjects) is available on neurovault (https://neurovault.org/collections/JBICXOQC/) [*to reviewers/editor: private collection, will be made public upon acceptance of manuscript*].

## 3. Results

### 3.1 EEG-informed fMRI analysis

#### Amyg-EFP-related brain-BOLD activation

Cluster-based control of type-I error resulted in 5 significant voxel-clusters, with one cluster including the right amygdala (amygdala-voxel with highest activation at [30,4,-19], MNI coordinates, T(15)=4.02). In line with a-priori expectations, we found that the correlation between Amyg-EFP and the effect size of fMRI-BOLD activation was replicated in large parts of the brain, including the amygdala (Figure 1).

**Figure 1.**
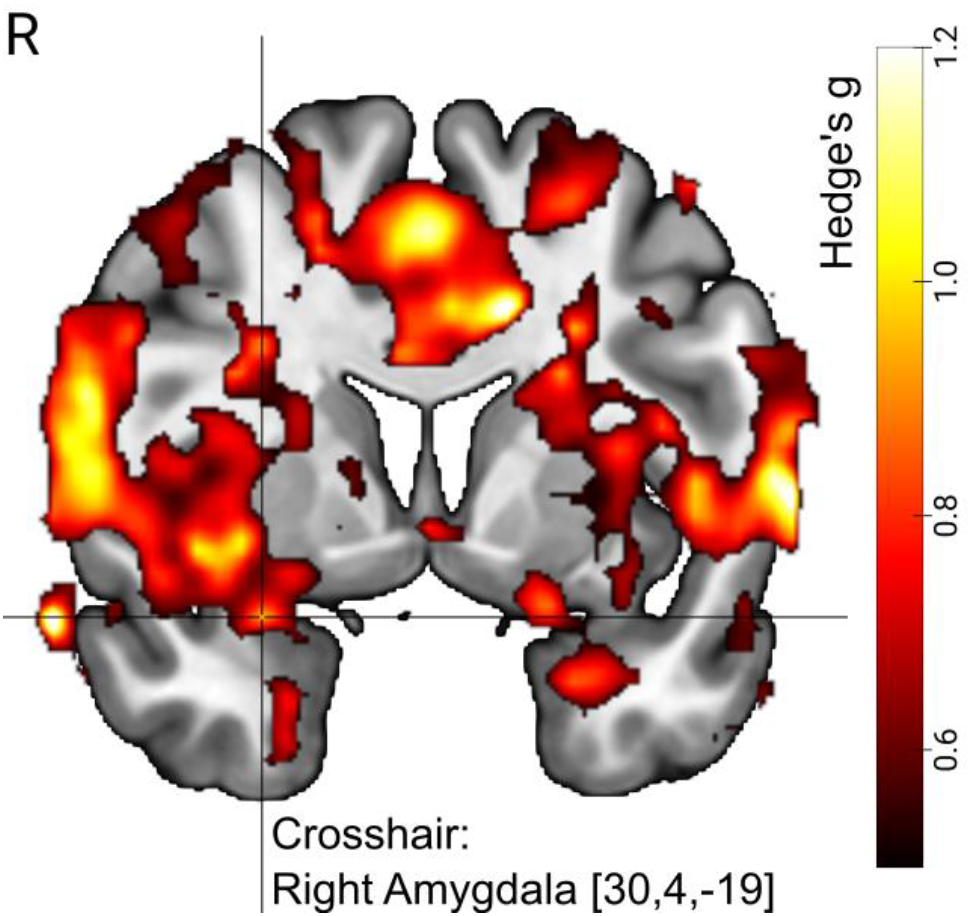
Amyg-EFP signal predicted right amygdala BOLD activation in N=16 individuals with Borderline Personality Disorder (BPD) undergoing simultaneous fMRI-EEG measurements. Visualization shows map of effect sizes (Hedge’s g). The BPD dataset served as the “replication sample” in the analysis to replicate previous findings from Keynan et al. (2016), i.e., the “reference sample”. Voxels shown are limited to those voxels with effect sizes that were within the 90%-CI of the reference sample. With other words, the image illustrates replicated effects. The visualization is further limited to voxels with medium effect size or higher (Hedge’s g>0.5). Crosshair position (MNI coordinates) indicates the amygdala region that was part of a significant cluster with size k=129,570 voxels. For significance testing we used cluster correction for multiple comparisons (p<0.05 FWE, k>149) with a cluster-defining threshold of p<0.001 (T(15)>3.728). R=right.

#### Exploring neurocircuitry engagement

In order to investigate the involvement of different neurocircuitries in the generation of the Amyg-EFP signal, we calculated correlation coefficients of the Amyg-EFP with different brain regions. The results show that the Amyg-EFP is positively correlated with regions of the negative affect neurocircuitry, the salience neurocircuitry and the cognitive control neurocircuitry. This finding was driven by significant correlation of Amyg-EFP with the BOLD-activation of bilateral amygdala, dorsal anterior cingulate cortex (dACC) and left dorsolateral prefrontal cortex (dlPFC, Figure 2). Note that some regions (including amygdala) are part of more than one circuitry. No significant correlations were observed with the DMN and the positive affect neurocircuitry. Correlations with visual, auditory and motor areas were significant and, descriptively, larger in comparison to the other networks. Due to the explorative nature of this analysis, we did not adjust significance thresholds to control for type-I error.

**Figure 2.**
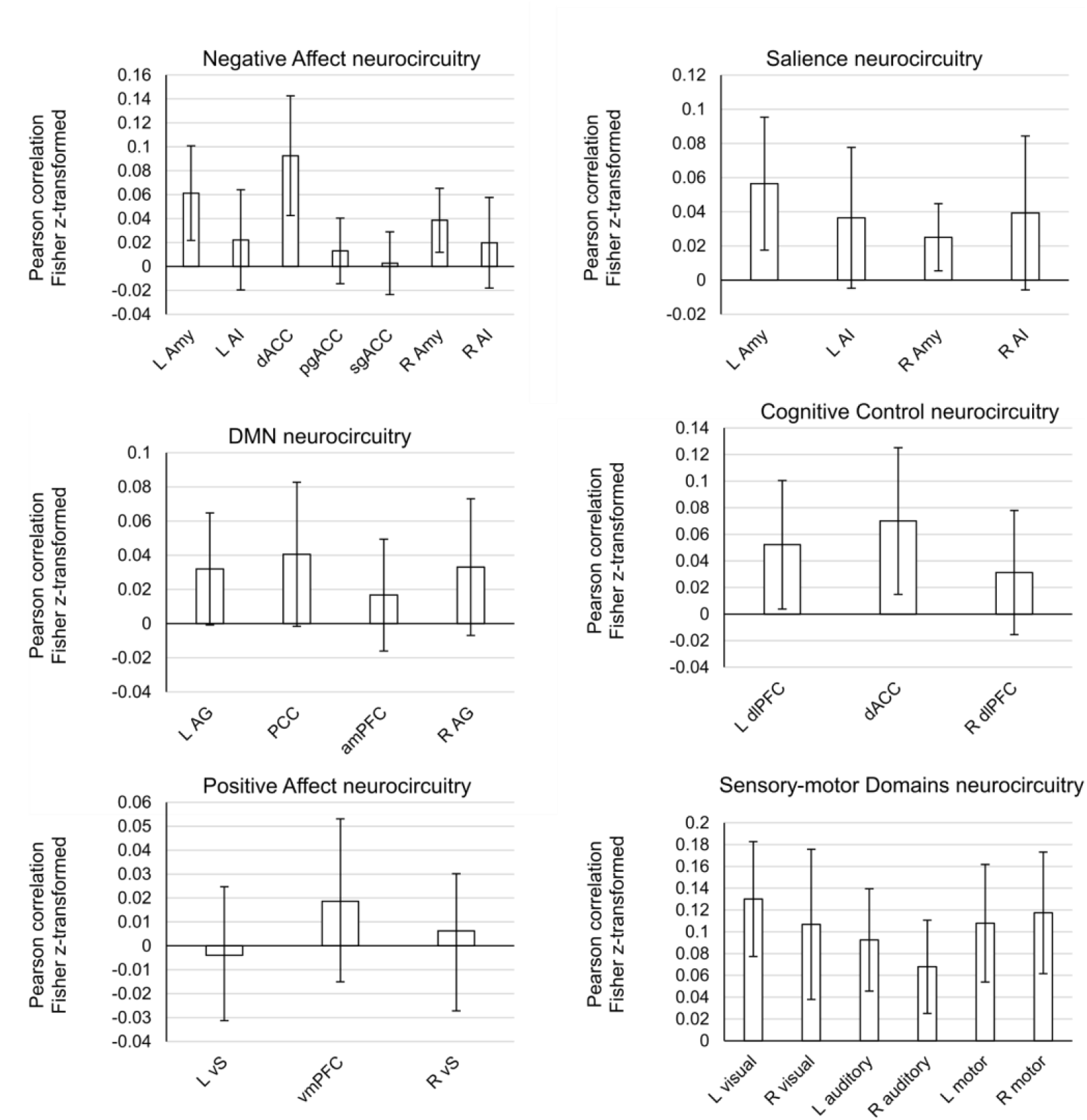
Correlations of Amyg-EFP with brain regions from different neurocircuitries (N=16). Mean Fisher-z transformed Pearson correlation is shown with 95% CI. Correlations can be said to be significant when the 95%-CI does not overlap with 0. Note that size of CIs was not corrected for multiple comparisons, limiting the utility of significance thresholds. L=left, R=right. Amy=Amygdala, AI=anterior insula, ACC=anterior cingulate cortex, dACC=dorsal ACC, pgACC=perigenual ACC, sgACC=subgenual ACC, AG=angular gyrus, PCC=posterior cingulate cortex, PFC=prefrontal cortex, amPFC=anterior medial PFC, dlPFC=dorsolateral PFC, vS=ventral striatum, vmPFC=ventromedial PFC.

### 3.2 Feasibility of Amyg-EFP training

#### High patient drop-out due to early discharge from residential DBT-program

Out of 51 patients (29 [number in brackets: NF-group participants]) entering the trial, 31 (15) completed the study until the post-measurement. This corresponds to a drop-out rate of 39% (48%). 12 (8) patients dropped out, because they left the residential DBT-program early.

When drop-outs due to early DBT termination are discounted, the drop-out rate is reduced to 16% (21%) (see Figure S1).

#### Clinical outcomes improved, no additive effect of NF was observed

Questionnaire data from the post-assessment of one subject was lost. After excluding an extreme value from the BDI-data analysis, we observed a non-significant Group main effect (ME) (F(23)=1.61, p=0.217), a significant Time ME (F(23)=8.84, p<0.01) indicating decreasing depression, and a non-significant Group x Time interaction (F(23)=2.87, p=0.104). The ANOVA of ALS scores showed a non-significant Group ME (F(24)=0.21, p=0.653), a significant Time ME (F(24)=7.78, p=0.01) of decreasing affective lability, and a non-significant Group x Time interaction (F(24)=0.30, p=0.591). No effects were significant in the ANOVA of TAS scores (Group ME: F(24)=2.42, p=0.133; Time ME: F(24)=3.80, p=0.063, Group x Time interaction (F(24)=0.33, p=0.572).

#### Patients improved regulation of the Amyg-EFP signal

15 participants completed the training regimen and entered the analysis. The analysis of Amyg-EFP NF training success resulted in a significant Session fixed effect (T(14.172)=2.375, p<0.05) and evidenced a linear increase of the Amyg-EFP as indicated by the PES measure across training sessions. The Subject intercept was significant (T(14.537)=3.624, p<0.005). Model fit was assessed with conditional *R*^*2*^_*GLMM*_=0.3545 (i.e., variance explained by the entire model) and marginal *R*^*2*^_*GLMM*_=0.042 (i.e., variance explained by fixed effects, Supplementary Figure S3). The analysis was repeated with the feedback values, i.e., the volume of the auditory interface, which is given as a number between 0=minimal and 1=maximal amplitude. Results of this analysis were consistent with the results from the previous analysis (Session fixed effect: T(14.150)=2.323, p<0.05, Subject intercept: T(14.955)=45.872, p<0.001; conditional *R*^*2*^_*GLMM*_=0.249, marginal *R*^*2*^_*GLMM*_=0.033) and showed, that subjects downregulated the auditory feedback in line with instructions (Figure 3).

**Figure 3.**
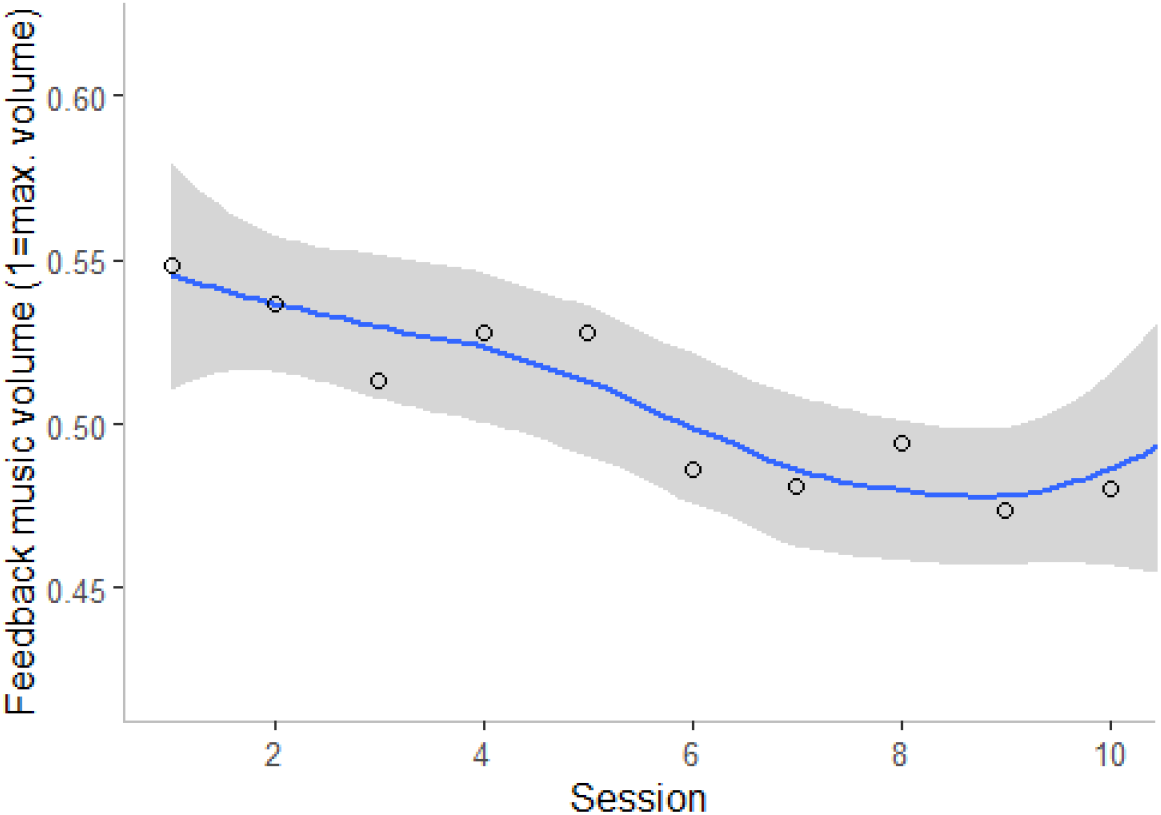
Participants learned to regulate the Amyg-EFP with neurofeedback training. N=15. Regression line with standard error is shown. Circles indicate session mean.

In line with reporting guidelines we explored correlations between NF training success (PES) and clinical improvement (N=14; affective lability: r=0.281, depression: r=0.236, alexithymia r=0.123; Online Supplement, Figure S3). Correlations were not significant.

## 4 Discussion

Borderline Personality Disorder (BPD) is associated with disturbed amygdala function and the modulation of the amygdala may be of therapeutic benefit for patients (Sicorello & Schmahl, 2021). Neurofeedback (NF) is a training based on reinforcement learning, empowering individuals to regulate brain activation based on feedback of brain activation (Lubianiker et al., 2022). Individuals can learn to self-modulate amygdala activation through NF, and clinical improvement with amygdala-targeted NF has been observed across different patient populations suffering from emotion dysregulation (Goldway et al., 2022). Previous proof-of-concept studies with individuals with BPD investigated amygdala self-regulation using functional Magnetic Resonance Imaging (fMRI)-based NF (Paret et al., 2016; Zaehringer et al., 2019). This study applied the Amyg-EFP, an EEG model of fMRI-defined amygdala activity, to BPD patients in order to test its predictive reliability and the feasibility of NF training in this cohort. Significance testing as well as formal replication analysis of whole-brain effect sizes confirmed correlation of the Amyg-EFP with deep-brain fMRI-Blood Oxygenation Level Dependent (BOLD) activation as hypothesized. These findings demonstrate that the Amyg-EFP is a generic EEG-based model of fMRI-BOLD activation that generalizes to clinical populations. Comparing effect sizes between two datasets, as performed in this analysis, enables replication analysis without the decision to exclude voxels based on conventional and somewhat arbitrary significance thresholds. Namely, the comparison of effect sizes can be regarded as a quantitative evaluation of replication success, as opposed to the visual, hence qualitative, evaluation of two significance maps from two independent analyses. Next, we assessed the feasibility of Amyg-EFP NF as an additional training during a 12-weeks residential Dialectical Behavior Therapy (DBT) program. Patients were able to improve regulation of the Amyg-EFP across 10 sessions. This finding aligns with previous research, showing that subjects can learn to modulate the Amyg-EFP with NF (Fruchtman-Steinbok et al., 2021; Goldway et al., 2018; Keynan et al., 2016, 2019). Since this study lacks an active control group, no conclusion can be drawn regarding specific treatment effects on the improved ability to regulate the Amyg-EFP-signal. A qualitative review of patient drop-out evidenced high drop-out rates in the overall sample as compared to rates usually observed in clinical trials (Dixon & Linardon, 2020). Increased drop-out was considerably driven by patients leaving the residential DBT treatment earlier than planned, due to reasons independent of this study. Discharge before the post-measurement led to exclusion from the study and inflated the drop-out rate of this trial.

Exploratory correlation analysis of the Amyg-EFP with regions of interest besides the amygdala revealed correlations with the dorsal anterior cingulate (dACC) and the dorsolateral prefrontal cortex (dlPFC). Furthermore, the Amyg-EFP signal mapped to the negative affect neurocircuitry, salience neurocircuitry, and cognitive control neurocircuitry, which relate to neurobehavioral functional domains as defined by the Research Domain Criteria (RDoC) (Goldstein-Piekarski et al., 2022; Insel et al., 2010). Additionally, activation from sensory and motor cortex correlated with the Amyg-EFP. Future studies comparing the Amyg-EFP to other e.g. more local brain measures are needed to investigate advantages and disadvantages of more locally focused brain signals for NF-training. A limitation concerns the explorative nature of this analysis. Hence, these findings require replication in an independent sample.

Originally, this study was designed to assess whether patients would generalize Amyg-EFP NF to amygdala-BOLD downregulation and whether the treatment would return a clinical benefit. A programming error in the NF-training script resulted in patients receiving feedback inverse to the original design defined in the trial protocol, i.e., Amyg-EFP upregulation instead of Amyg-EFP downregulation was rewarded with positive feedback for the participant. This programming error was only detected after data collection was completed.

Therefore, it is not possible to assess generalization and clinical utility with this dataset. We did not observe any differences between the NF group and the DBT-only group in clinical measures, and no correlations of NF success and clinical change were significant, suggesting that patients incurred no harm from upregulating Amyg-EFP. While being an accidental finding, this preliminary evidence indicates that upregulation might be a safe control condition in future studies to assess causality of directional control of brain activation and clinical improvement (called ‘bidirectional-regulation’ control condition by Sorger and colleagues (2019)). Future research confirming low risk and clinical trials implementing bidirectional control with Amyg-EFP NF can be of high value in face of the multiple mechanisms driving the effects of NF training (Ros et al., 2020).

### 4.1 Conclusions

The Amyg-EFP correlates with deep-brain activation and can be modulated with neurofeedback training. This research extends previous findings to a different laboratory and to a different sample, namely female patients with Borderline Personality Disorder.

Salience/negative affect neurocircuitry and cognitive control neurocircuitry activation was found to correlate with the Amyg-EFP, a finding that awaits replication in future studies. Neurofeedback as add-on therapeutic training is feasible, although considerable patient dropout in the residential treatment context was observed. Future research investigating the clinical utility of Amyg-EFP neurofeedback in BPD is needed.

## Data Availability

Self-report data, individual fMRI and EEG data is available on reasonable request and can be shared in line with applicable data protection regulations. Analysis code of self-report and NF training data is openly available. The T-map from the second-level fMRI analysis (i.e., aggregated data across subjects) is available on neurovault.

https://doi.org/10.5281/zenodo.7766644

https://neurovault.org/collections/JBICXOQC/

## 4.2 CRediT authorship contribution statement

CS: Conceptualization, Supervision, Writing - review & editing

CP: Conceptualization, Methodology, Data Curation, Formal analysis, Verification, Supervision, Visualization, Writing - Original Draft.

GG, JNK: Software, Data Curation, Formal analysis, Writing - review & editing.

MJ: Investigation, Writing - review & editing

MZ: Investigation, Project administration, Data Curation, Formal analysis, Visualization, Writing - Original Draft

SB, PA, DB: Methodology (EEG), Writing - review & editing

SC: Conceptualization, Investigation

TH: Conceptualization, Supervision, Software, Writing - review & editing.

## 4.4 Acknowledgement

The authors want to thank Madita Stirner for conducting patient measurements, Nike Unverhau and Ida Mueller for contributing R-code, Florian Kirn for R-code review, Maurizio Sicorello for support with MLM analysis, and Martin Fungisai Gerchen for helpful advice on fMRI-replicability analysis.

## Footnotes

1 Online preregistration: https://doi.org/10.17605/OSF.IO/6ZDS5, clinicaltrials.org: NCT03964545

