## Supplement for "Amygdala-related electrical fingerprint is modulated with neurofeedback training and correlates with deep-brain activation: Proof-of-concept in borderline personality disorder"

### List of Content

**List of Content** 2
**1 Supplementary Methods** 3
1.1 MRI-EEG procedure and experimental task 3
1.2 Preprocessing of fMRI data 3
1.3 Real-time fMRI analysis and amygdala BOLD-feedback computation 7
**2 Supplementary Figures** 9
**3 Supplementary Tables** 13
**4 References** 14

### 1 Supplementary Methods

#### 1.1 Simultaneous fMRI-EEG procedure and experimental task

After positioning of the EEG cap and impedance check, participants entered the MR scanner. Their heads were lightly restrained in the head coil to limit movement during scanning. A computer screen was visible via a mirror placed upon the head coil. Impedances of EEG electrodes were checked before the MR measurement started and the helium pump was switched off to reduce artifacts. Participants were instructed to lie still in the scanner with eyes open and look at a fixation cross displayed on a blank computer screen during the resting state scan. During the neurofeedback (nf) runs, participants were instructed to downregulate a thermometer displayed on the screen. Each nf block consisted of a rest phase, followed by amygdala feedback and finger-tapping task as illustrated in Figure S1.

#### 1.2 Preprocessing of fMRI data

Results included in this manuscript come from preprocessing performed using *fMRIPrep* 20.0.6 (Esteban, Markiewicz, et al. (2018); Esteban, Blair, et al. (2018); RRID:SCR_016216), which is based on *Nipype* 1.4.2 (Gorgolewski et al. (2011); Gorgolewski et al. (2018); RRID:SCR_002502).

**Anatomical data preprocessing**

A total of 2 T1-weighted (T1w) images were found within the input BIDS dataset. All of them were corrected for intensity non-uniformity (INU) with N4BiasFieldCorrection (Tustison et al. 2010), distributed with ANTs 2.2.0 (Avants et al. 2008, RRID:SCR_004757). The T1w-reference was then skull-stripped with a *Nipype* implementation of the antsBrainExtraction.sh workflow (from ANTs), using OASIS30ANTs as target template. Brain tissue segmentation of cerebrospinal fluid (CSF), white-matter (WM) and gray-matter (GM) was performed on the brain-extracted T1w using fast (FSL 5.0.9, RRID:SCR_002823, Zhang, Brady, and Smith 2001). A T1w-reference map was computed after registration of 2 T1w images (after INU-correction) using mri_robust_template (FreeSurfer 6.0.1, Reuter, Rosas, and Fischl 2010). Brain surfaces were reconstructed using recon-all (FreeSurfer 6.0.1, RRID:SCR_001847, Dale, Fischl, and Sereno 1999), and the brain mask estimated previously was refined with a custom variation of the method to reconcile ANTs-derived and FreeSurfer-derived segmentations of the cortical gray-matter of Mindboggle (RRID:SCR_002438, Klein et al. 2017). Volume-based spatial normalization to one standard space (MNI152NLin2009cAsym) was performed through nonlinear registration with antsRegistration (ANTs 2.2.0), using brain-extracted versions of both T1w reference and the T1w template. The following template was selected for spatial normalization: *ICBM 152 Nonlinear Asymmetrical template version 2009c* [Fonov et al. (2009), RRID:SCR_008796; TemplateFlow ID: MNI152NLin2009cAsym],

**Functional data preprocessing**

For each of the 5 BOLD runs found per subject (across all tasks and sessions), the following preprocessing was performed. First, a reference volume and its skull-stripped version were generated using a custom methodology of *fMRIPrep*. A B0-nonuniformity map (or *fieldmap*) was estimated based on a phase-difference map calculated with a dual-echo GRE (gradient-recall echo) sequence, processed with a custom workflow of *SDCFlows* inspired by the epidewarp.fsl script and further improvements in HCP Pipelines (Glasser et al. 2013). The *fieldmap* was then co-registered to the target EPI (echo-planar imaging) reference run and converted to a displacements field map (amenable to registration tools such as ANTs) with FSL's fugue and other *SDCflows* tools. Based on the estimated susceptibility distortion, a corrected EPI (echo-planar imaging) reference was calculated for a more accurate co-registration with the anatomical reference. The BOLD reference was then co-registered to the T1w reference using bbregister (FreeSurfer) which implements boundary-based registration (Greve and Fischl 2009). Co-registration was configured with six degrees of freedom. Head-motion parameters with respect to the BOLD reference (transformation matrices, and six corresponding rotation and translation parameters) are estimated before any spatiotemporal filtering using mcflirt (FSL 5.0.9, Jenkinson et al. 2002). BOLD runs were slice-time corrected using 3dTshift from AFNI 20160207 (Cox and Hyde 1997, RRID:SCR_005927). The BOLD time-series (including slice-timing correction when applied) were resampled onto their original, native space by applying a single, composite transform to correct for head-motion and susceptibility distortions. These resampled BOLD time-series will be referred to as *preprocessed BOLD in original space*, or just *preprocessed BOLD*. The BOLD time-series were resampled into standard space, generating a *preprocessed BOLD run in MNI152NLin2009cAsym space*. First, a reference volume and its skull-stripped version were generated using a custom methodology of *fMRIPrep*. Several confounding time-series were calculated based on the *preprocessed BOLD*: framewise displacement (FD), DVARS and three region-wise global signals. FD and DVARS are calculated for each functional run, both using their implementations in *Nipype* (following the definitions by Power et al. 2014). The three global signals are extracted within the CSF, the WM, and the whole-brain masks. Additionally, a set of physiological regressors were extracted to allow for component-based noise correction (*CompCor*, Behzadi et al. 2007). Principal components are estimated after high-pass filtering the *preprocessed BOLD* time-series (using a discrete cosine filter with 128s cut-off) for the two *CompCor* variants: temporal (tCompCor) and anatomical (aCompCor). tCompCor components are then calculated from the top 5% variable voxels within a mask covering the subcortical regions. This subcortical mask is obtained by heavily eroding the brain mask, which ensures it does not include cortical GM regions. For aCompCor, components are calculated within the intersection of the aforementioned mask and the union of CSF and WM masks calculated in T1w space, after their projection to the native space of each functional run (using the inverse BOLD-to-T1w transformation). Components are also calculated separately within the WM and CSF masks. For each CompCor decomposition, the *k* components with the largest singular values are retained, such that the retained components' time series are sufficient to explain 50 percent of variance across the nuisance mask (CSF, WM, combined, or temporal). The remaining components are dropped from consideration. The head-motion estimates calculated in the correction step were also placed within the corresponding confounds file. The confound time series derived from head motion estimates and global signals were expanded with the inclusion of temporal derivatives and quadratic terms for each (Satterthwaite et al. 2013). Frames that exceeded a threshold of 0.5 mm FD or 1.5 standardised DVARS were annotated as motion outliers. All resamplings can be performed with *a single interpolation step* by composing all the pertinent transformations (i.e. head-motion transform matrices, susceptibility distortion correction when available, and co-registrations to anatomical and output spaces). Gridded (volumetric) resamplings were performed using antsApplyTransforms (ANTs), configured with Lanczos interpolation to minimize the smoothing effects of other kernels (Lanczos 1964). Non-gridded (surface) resamplings were performed using mri_vol2surf (FreeSurfer).

Many internal operations of *fMRIPrep* use *Nilearn* 0.6.2 (Abraham et al. 2014, RRID:SCR_001362), mostly within the functional processing workflow. For more details of the pipeline, see the section corresponding to workflows in *fMRIPrep*'s documentation.

**Copyright Waiver**

The above boilerplate text was automatically generated by fMRIPrep with the express intention that users should copy and paste this text into their manuscripts *unchanged*. It is released under the CC0 license.

#### 1.3 Real-time fMRI analysis and amygdala BOLD-feedback computation

**Localization of the amygdala.** The brain mask definition, the quantification of BOLD-signal and the neurofeedback presentation to the participant was implemented as reported in a prior study by our group (Paret et al., 2018). Following image reconstruction at the scanner, volumes were transferred to a laptop for preprocessing and analysis with SPM12 (Wellcome Department of Cognitive Neurology, London, UK). Before beginning the neurofeedback tasks, the T1-weighted anatomical scan was segmented and normalized to Montreal Neurological Imaging (MNI) space. Then the anatomical masks of the Regions of Interest (ROIs) for feedback calculation were transformed to subject-native space. The ROI in this study was the right amygdala mask, produced with the Harvard–Oxford brain atlas with a probability threshold of 25%. For each participant, BOLD signal from the voxels within this mask were used to calculate their neurofeedback values. In order to correct for global signal fluctuations across the brain, the BOLD signal from a rectangular plane ROI (3x30x30 mm in AC–PC orientation, center of mass = [0,216,25], MNI coordinates) was recorded and subtracted from the target-ROI BOLD-activation.

**Quantification of BOLD signals for neurofeedback calculation.** Functional images were realigned to the first image. For each ROI, BOLD signal data from all voxels were averaged. The resulting average time course was processed with a modified Kalman filter (Koush et al.,2012) and then detrended with Matlab’s (R2014b) detrend function. Detrending started with the 35th acquired volume before subjects received any feedback to allow for stabilization of the filter and detrend functions. Following this signal preprocessing, percent signal change from the global mean was calculated. The difference in signal change between the ROIs was used for feedback:

$$Feedback_i = detrend(filter(\frac{X_{amygdala,i} * 100}{mean(X_{amygdala,\, 1 \rightarrow i})})) - detrend(filter(\frac{X_{control,i} * 100}{mean(X_{control,\, 1 \rightarrow i})}))$$

where $X_{j,i}$ is the most recent BOLD signal value received at the ROI.

**MR-BOLD feedback presentation.** Stimulus presentation software (Presentation, Neurobehavioral Systems, Berkeley, CA) running on a separate computer received the score data via TCP/IP. The feedback display was refreshed upon receiving the next score with an update frequency of 0.5 Hz. Feedback was displayed as a colored rectangle moving on a vertical thermometer-like scale. In addition to moving up and down, the rectangle changed from dark red at the top of the scale over light green in the middle to dark green at the bottom. The thermometer-like scale had a resolution of six levels and adjusted the rectangle's position within those levels according to variations within a range of two percent signal change above and below baseline.

### 2 Supplementary Figures

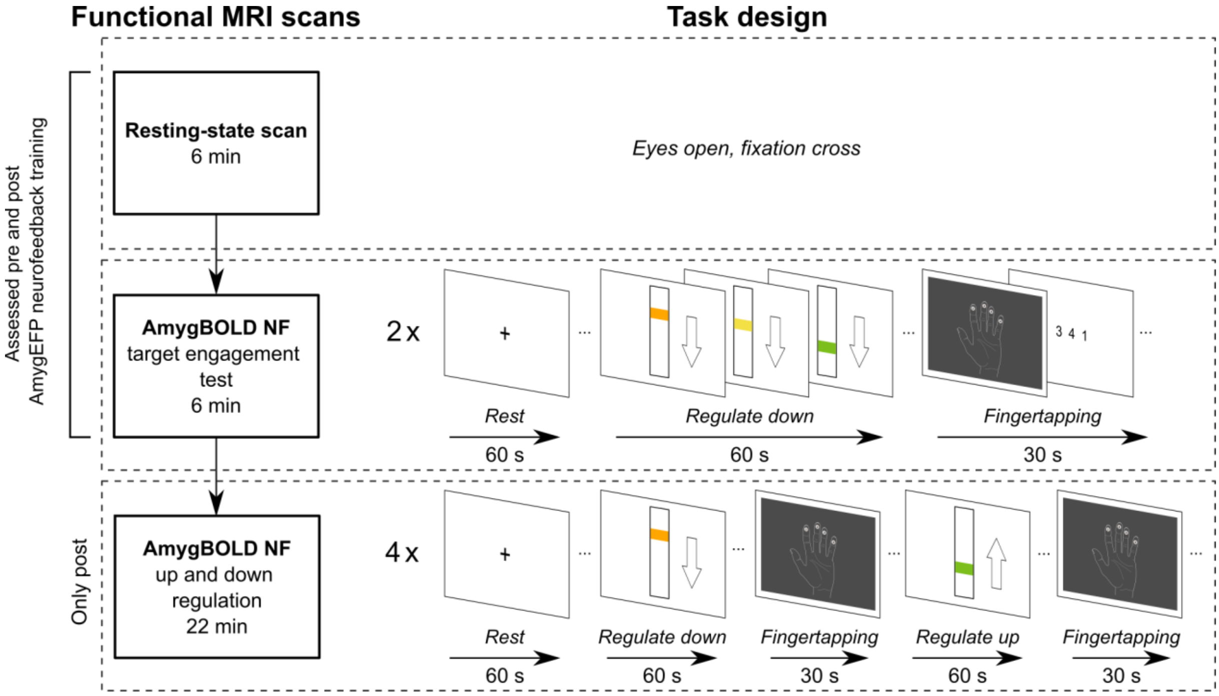

Supplementary Figure S1. Procedure of the fMRI-NF session at the beginning ("pre") and at the end ("post") of the study. Two different Amygdala-BOLD-NF tasks were performed: a brief 2-block downregulation-NF session (termed "EFPTest") and a longer 4-block NF-session including alternating up- or downregulation blocks (termed "UpDown")

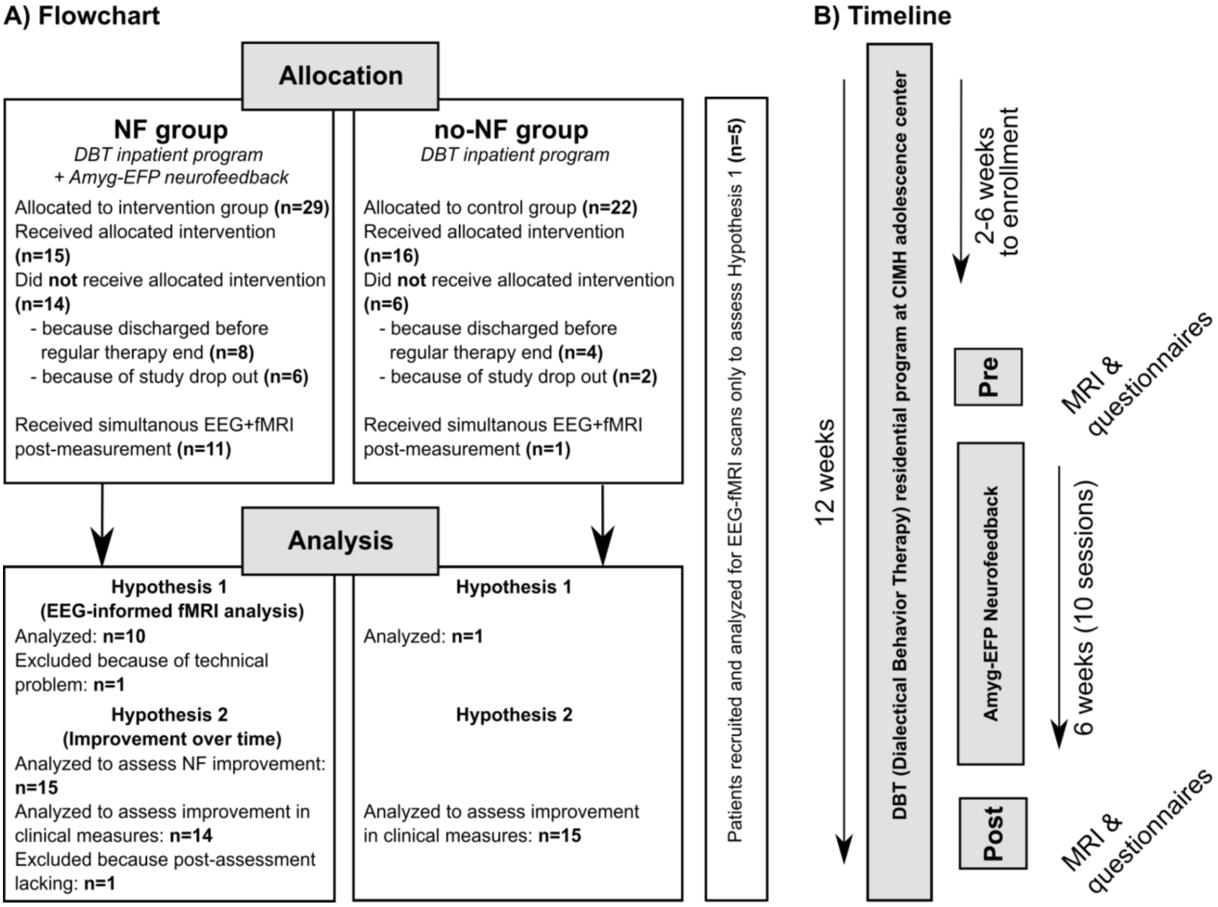

Supplementary Figure S2. A) Patient flow chart including drop-out numbers and reason for dropping out of the study. B) The study's timeline, which participants completed within the context of their 3-month residential therapy program.

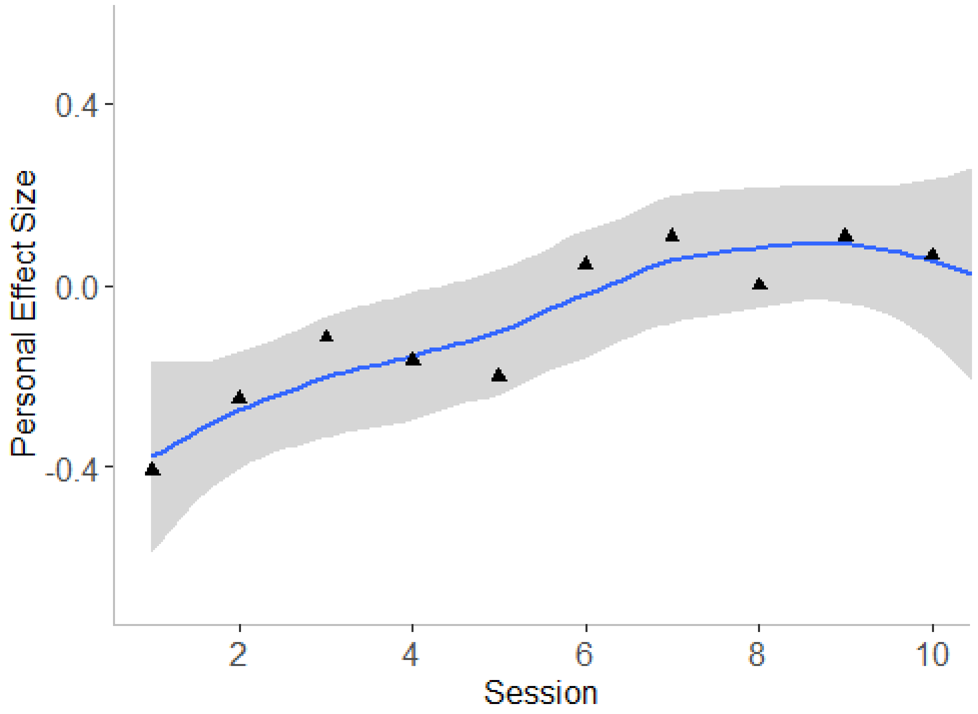

Supplementary Figure S3. Participants learned to regulate the Amyg-EFP with neurofeedback training. N=15. Regression line with standard error is shown. Triangles indicate session mean.

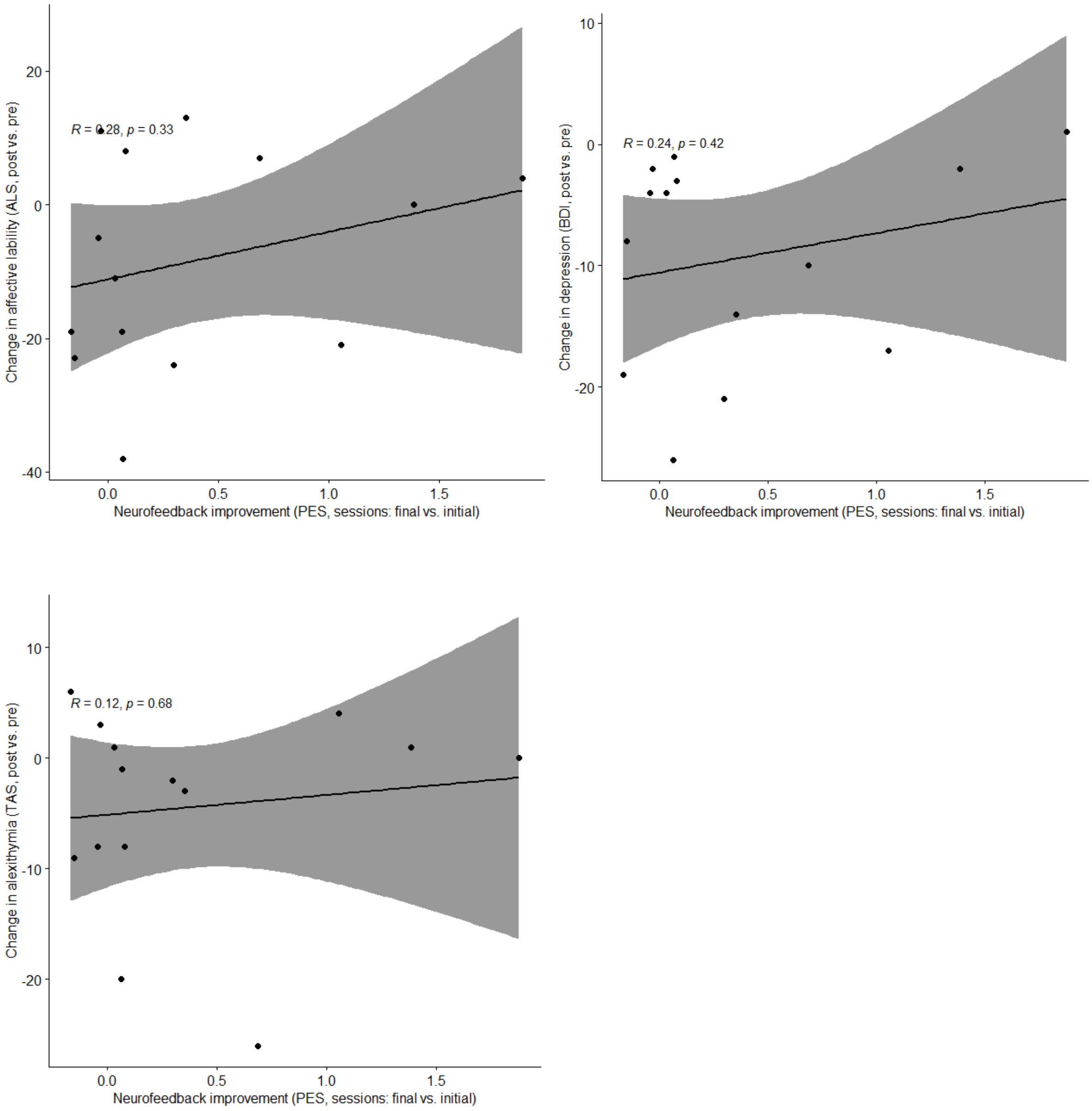

Supplementary Figure S4. Correlations of improvement in clinical outcomes (difference of post vs. pre) and neurofeedback training success (difference of means of final two and initial two sessions, N=14). Linear trend with 95%-confidence interval is shown. ALS=Affective Lability, BDI=Beck Depression Inventory, TAS=Toronto Alexithymia Scale, PES=personal effect size.

### 3 Supplementary Tables

Table S1: Replication (BPD) sample, study 1. Sample characteristics.

| | | |
|---|---|---|
| ***Demographics*** | | |
| *N* | 16 | |
| female sex N (%) | 16 | (100) |
| Age mean (SD) | 21.30 | (2,19) |
| ***Self-report measures*** | | |
| *N* | 15 | |
| ***Beck Depression Inventory (BDI)*** | Mean | SD |
| Total (SD) | 30.47 | 13.92 |
| ***Affect Lability Scale (ALS)*** | | |
| Total (SD) | 86.13 | 30.28 |
| Depression | 19.00 | 6.40 |
| Hypomania | 14.93 | 7.70 |
| Biphasic shifts | 12.87 | 5.82 |
| Anxiety | 12.93 | 4.38 |
| Anger | 9.33 | 7.07 |
| Anxiety Depression | 17.07 | 4.91 |
| ***Toronto Alexithymia Scale (TAS-26)*** | | |
| Total (SD) | 55.53 | 11.33 |
| Identification of one's feelings | 22.73 | 7.17 |
| Difficulty Describing Feelings | 19.13 | 3.94 |
| External thinking | 13.67 | 3.81 |

Table S2. CRED-nf checklist

*Will be provided following formal acceptance of the manuscript by journal.*
